# Evaluating the quality of prostate cancer diagnosis recording in routinely collected primary care data for observational research: A study using multiple linked English electronic health records databases

**DOI:** 10.1101/2024.08.21.24312333

**Authors:** Gayasha Somathilake, Elizabeth Ford, Jo Armes, Sotiris Moschoyiannis, Michelle Collins, Patrick Francsics, Agnieszka Lemanska

## Abstract

**Background:** Primary care data in the UK are widely used for cancer research, but the reliability of recording key events such as diagnoses remains uncertain. Data linkage can mitigate these uncertainties; however, researchers may avoid linkage due to high costs, tight timelines, and sample size limitations. Hence, this study aimed to assess the quality of prostate cancer (PCa) diagnoses in primary care. We utilised Clinical Practice Research Datalink (CPRD) primary care data linked to National Cancer Registration and Analysis Service (NCRAS) and Hospital Episode Statistics (HES) in England. We compared accuracy, completeness, and timing of diagnosis recording between sources to facilitate decision-making regarding data source selection for future research.

**Methods:** Incident PCa diagnoses (2000-2016) for males aged ≥46 years recorded in at least one study data source were examined. The accuracy of a data source was estimated by the proportion of diagnoses recorded in the specific source that was also confirmed by any linked source. Completeness was estimated by identifying the proportion of all diagnoses in linked sources with a matching diagnosis in the specific source.

**Results:** The study included 51,487 PCa patients from either source. CPRD demonstrated 86.9% accuracy and 68.2% completeness against NCRAS and 75.1% accuracy and 61.1% completeness against HES. Overall, CPRD showed the highest accuracy (93%) but the lowest completeness (60.7%). Diagnosis dates in CPRD were more concordant with NCRAS (90.6% within 6 months) than with HES (61.2%). Over time, accuracy and completeness improved, especially after 2004. Discrepancies in diagnosis dates revealed a median delay of 2 weeks in CPRD than NCRAS and 1 week than HES. CPRD Aurum exhibited better quality compared to GOLD.

**Conclusions:** While the accuracy of PCa diagnoses in CPRD compared to linked sources was high, completeness was low. Therefore, linking to HES or NCRAS should be considered for improved case capture, acknowledging their inherent limitations.

## 1. Introduction

In the United Kingdom (UK), primary care electronic health record (EHR) databases cover over 98% of the population and are extensively used for cancer research since most patients with symptoms initially present to a general practitioner (GP) (1–5). However, identifying cancer diagnoses from primary care EHRs presents challenges due to inaccurate and incomplete data varying across cancer types (6–8). Additionally, a diagnostic code in primary care may not definitively indicate a cancer diagnosis, while lacking a code may not consistently imply the absence of a diagnosis (9). Recent improvements in data linkage provide solutions to mitigate these uncertainties and the potential misclassification bias when studies solely rely on primary care data (10). Nonetheless, researchers might still avoid data linkage due to high costs, tight timelines, and limitations in sample size and coverage (11). This underscores the importance of assessing the correctness, completeness, and timeliness of primary care data for research purposes (12).

Prostate cancer (PCa) is the most common malignancy among men in the UK typically diagnosed with GP referrals following abnormal exams or elevated prostate-specific antigen levels. (13,14). For diagnosis information to appear in primary care records, hospital discharge or diagnosis letters must be accurately coded into EHRs. This can cause delays, as primary care diagnosis records are often only updated when GPs take action, such as prescribing medication (15). Moreover, errors in data transfer, or misclassification, such as confusing benign prostatic hyperplasia with PCa make identifying PCa diagnoses from primary care EHRs challenging. Hence, this study aimed to evaluate the quality of PCa diagnosis recording in primary care using linked Clinical Practice Research Datalink (CPRD) data in England.

CPRD is a primary care database with data from over 2,200 general practices, covering more than 60 million patients in the UK (16–18). The data available in two databases GOLD and Aurum can be linked to National Cancer Registration and Analysis Service (NCRAS), Hospital Episode Statistics (HES), and Office of National Statistics (ONS) (17). The study evaluated the quality of PCa diagnoses in CPRD based on accuracy (i.e. correctness), completeness (i.e. presence/missingness) and recording dates compared to NCRAS and HES (19). We also compared the recording quality between GOLD and Aurum datasets and examined patient records with diagnoses in linked sources but without corresponding primary care records. The findings aim to provide insights into primary care data quality and facilitate decision-making in data source selection for PCa research.

## 2. Methods

### 2.1 Data sources

This study utilised CPRD GOLD and Aurum data (November 2019 release version, set 17), linked to NCRAS, HES, ONS and 2015 Index of Multiple Deprivation (IMD) quintiles (20). The study was approved by the CPRD Independent Scientific Advisory Committee (protocol number 19_050R).

CPRD, one of the largest global EHR databases, contains de-identified longitudinal data from UK primary care practices, including demographics, diagnoses, prescriptions, tests, and referrals. It offers a secure data linkage service for external healthcare and area-level databases and is provided to a subset of English general practices that have given their consent and provided patient-level information. This linkage is established through a unique patient identifier created by CPRD, allowing for the integration and analysis of data from multiple sources (18,21).

NCRAS serves as the national standard for reporting cancer in England, incorporating data from the Cancer Registry (CR), Radiotherapy Dataset (RTDS), and Systemic Anti-Cancer Therapy (SACT). It is a dynamic population-based database that captures information across the entire cancer care pathway including cancer diagnoses, treatments, and outcomes. Cancer diagnoses are recorded using ICD-10 codes and the information is collected from hospitals, pathology and treatment reports, hospices, cancer screening and treatment centres (22).

HES records all admissions, outpatient appointments, and Accident and Emergency attendances at National Health Service (NHS) hospitals. HES Admitted Patient Care (APC) includes admission and discharge dates, diagnoses and procedures for inpatient hospitalisations. HES Outpatient (OP) data includes outpatient consultation details, appointment dates, specialty information, and clinical diagnoses and procedures (23).

### 2.2 Patient selection

The data extract included patients born in 1954 or earlier, with at least one year of continuous registration between 01/01/1990 and 31/12/2016, registered at English practices contributing data of acceptable quality to GOLD or Aurum and were linkable to NCRAS, HES, and IMD data. The patients from practices that switched from InPS Vision to EMIS were not included in the data extract to prevent duplication between GOLD and Aurum.

For this study, male patients with a record of a primary diagnosis of incident PCa between 01/01/2000 and 31/12/2016 were identified using predefined Read, SNOMED and ICD-10 diagnostic codes (24) (Supplementary_information-S1). The study period was decided when the data sources were concurrently available (Supplementary_information-S2). The date of the earliest diagnostic PCa code in any data source was assumed to be the diagnosis date. Active follow-up started on the latest registration date, up-to-standard date (when the general practice began providing continuous data), or the start of the study period. Follow-up ended at the earliest transfer out date, last data collection, death date, or the end of the study period. Patients with at least one year of continuous registration preceding the diagnosis dates within the active follow-up period were included.

### 2.3 Patient characteristics

We considered patient’s age in years at the time of diagnosis (<60, 60-69, 70-79, ≥80), the year of diagnosis (2000-2003, 2004-2007, 2008-2011, 2012-2016), the geographical region of the practice in England, the IMD quintile (1=least deprived to 5=most deprived), and the ethnicity of the patient (aggregated based on the higher-level ethnicity classification with six groups: Asian, Black, Mixed, Other, Unknown, White).

### 2.4 Data analysis

Henceforth, primary care data from CPRD GOLD or Aurum will be referred to as ‘CPRD,’ cancer registrations from CR, RTDS, and/or SACT as ‘NCRAS,’ and secondary care data from HES APC and/or OP as ‘HES.’ Comparisons between CPRD and NCRAS will be denoted as ‘CPRD-NCRAS’ and between CPRD and HES as ‘CPRD-HES. for ease of reference.

Accuracy and completeness were estimated using methodologies by Weiskopf and Weng (19). Accuracy of a study source was defined by the proportion of patients with PCa diagnoses in the specific source that had corresponding records in at least one linked source. Completeness of a source was defined as the proportion of all patients with diagnoses in linked sources that had corresponding records in the specific source. These metrics were calculated for CPRD-NCRAS and CPRD-HES and were further evaluated by the patient’s age, diagnosis year, IMD, and ethnicity.

Discrepancies in diagnosis dates were evaluated for CPRD-NCRAS and CPRD-HES considering diagnoses recorded within 6 months were indicative of the same cancer episode. Patients in CPRD who had corresponding diagnoses in NCRAS and/or HES recorded within 6 months were included in this analysis. PCa diagnosis recording was also compared between GOLD and Aurum to help inform decisions in selecting CPRD data for research purposes. Patients diagnosed in linked sources but without corresponding records in CPRD were further examined to elucidate potential reasons for missing primary care records.

This study adhered to the REporting of studies Conducted using the Observational Routinely collected health Data (RECORD) statement (25) for reporting observational research involving routinely collected data (Supplementary_information-S3). All statistical analyses were performed using R software (version 4.3.2).

## 3. Results

### 3.1 Study cohort

The final cohort included 51,487 patients with a PCa diagnosis recorded in at least one study source during follow-up (Fig. 1).

**Fig. 1.**
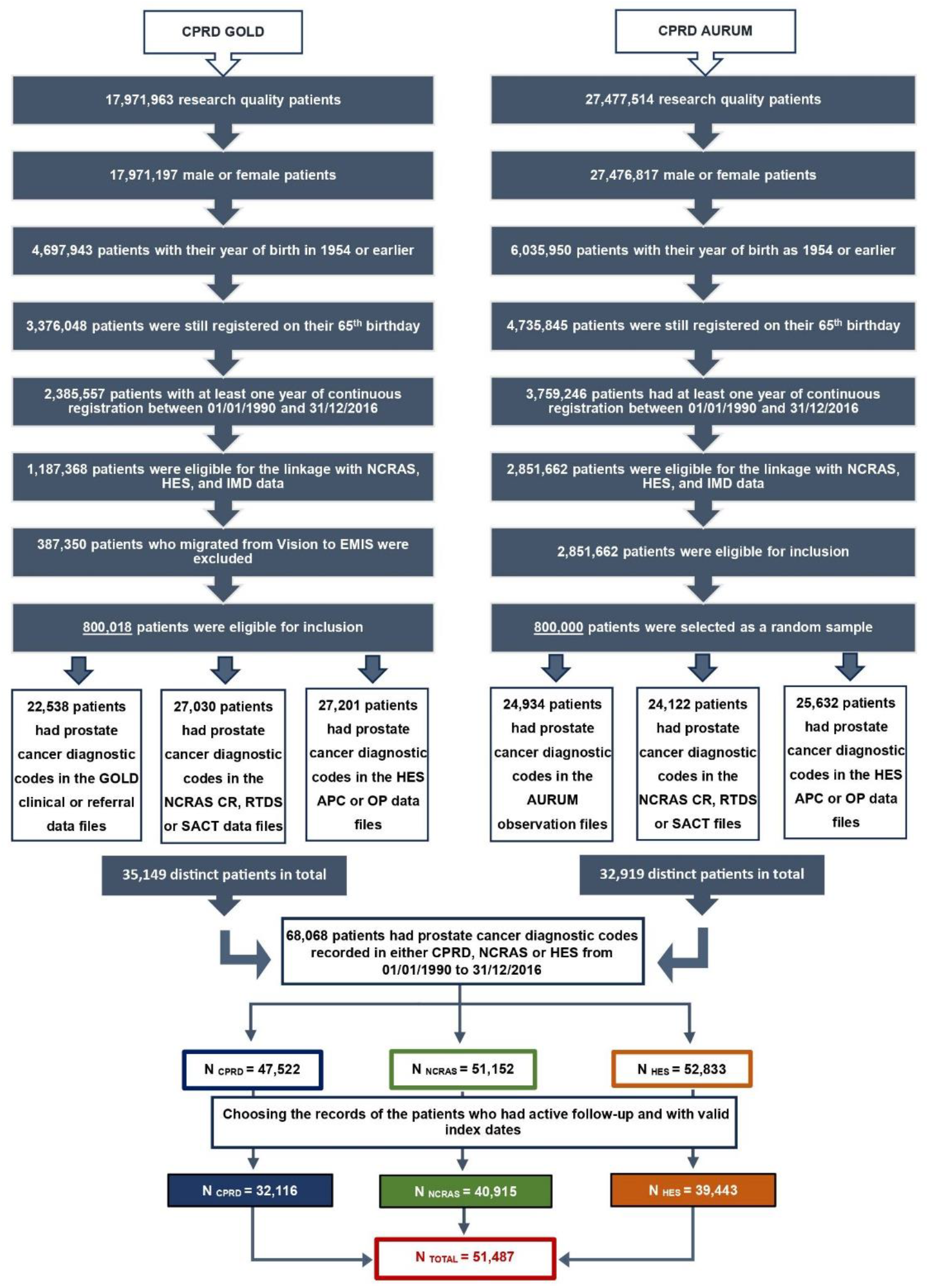
Workflow for extracting prostate cancer diagnoses from each study source

Age at diagnosis varied across sources, with HES indicating a higher proportion of patients aged ≥80 and fewer patients <60 compared to CPRD and NCRAS (Table 1). PCa cases increased over time in all sources, with CPRD compared to NCRAS and HES showing more diagnoses in earlier years (2000-2007) and fewer in later years (2012-2016). Most patients across all sources were from practices in the Southwest region. In CPRD, there was a higher proportion of cases in the IMD quintile 1 (least deprived) compared to NCRAS and HES, whereas the proportions for the IMD quintiles 4-5 (more deprived) were lower. Ethnicity was distributed similarly between the sources with the majority from the White ethnic group.

**Table 1.**
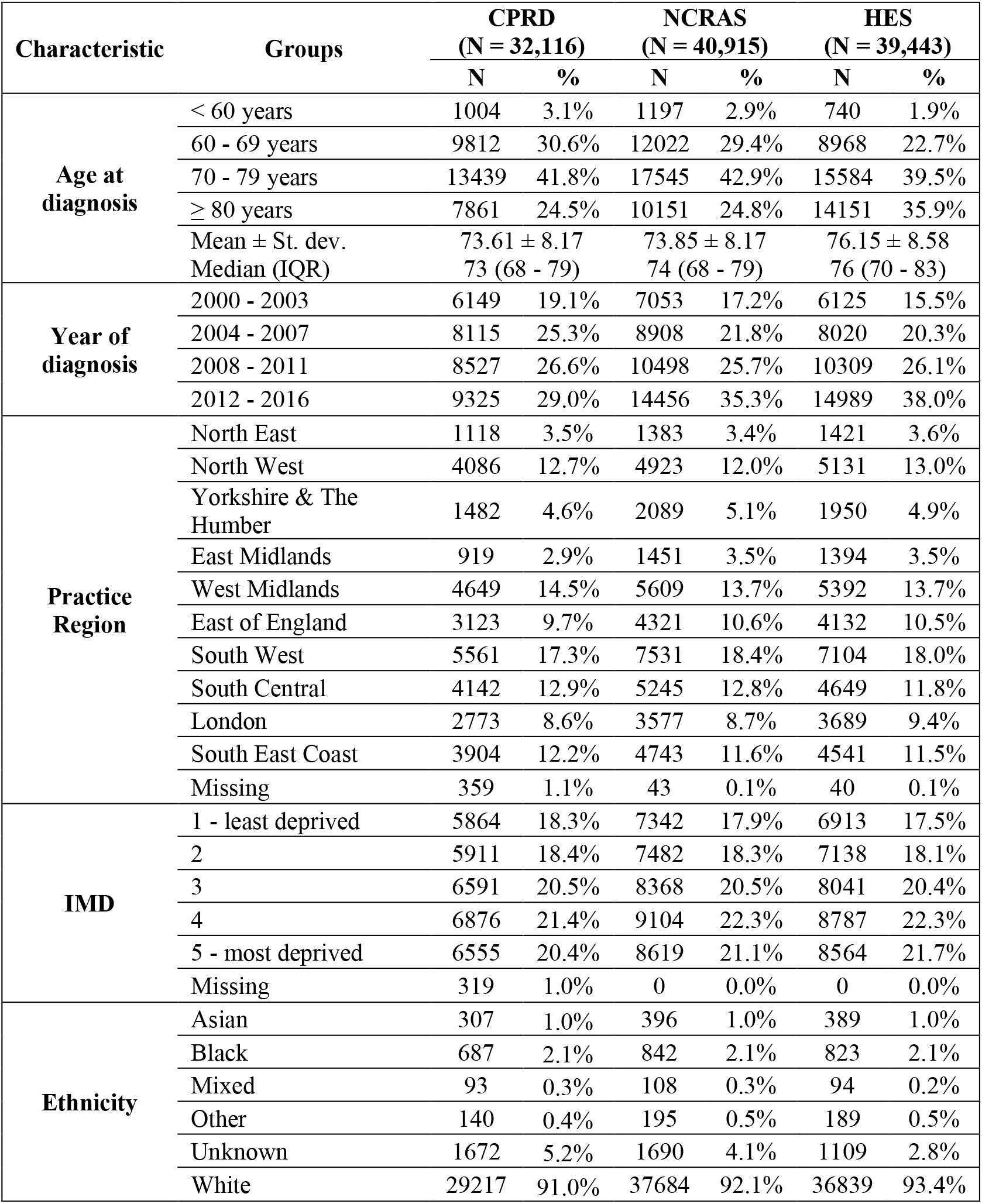
Patient characteristics between study data sources.

| Characteristic | Groups | CPRD<br>(N = 32,116) |  | NCRAS<br>(N = 40,915) |  | HES<br>(N = 39,443) |  |
| --- | --- | --- | --- | --- | --- | --- | --- |
|  |  | N | % | N | % | N | % |
| Age at diagnosis | < 60 years | 1004 | 3.1% | 1197 | 2.9% | 740 | 1.9% |
|  | 60 - 69 years | 9812 | 30.6% | 12022 | 29.4% | 8968 | 22.7% |
|  | 70 - 79 years | 13439 | 41.8% | 17545 | 42.9% | 15584 | 39.5% |
|  | ≥ 80 years | 7861 | 24.5% | 10151 | 24.8% | 14151 | 35.9% |
|  | Mean ± St. dev. | 73.61 ± 8.17 |  | 73.85 ± 8.17 |  | 76.15 ± 8.58 |  |
|  | Median (IQR) | 73 (68 - 79) |  | 74 (68 - 79) |  | 76 (70 - 83) |  |
| Year of diagnosis | 2000 - 2003 | 6149 | 19.1% | 7053 | 17.2% | 6125 | 15.5% |
|  | 2004 - 2007 | 8115 | 25.3% | 8908 | 21.8% | 8020 | 20.3% |
|  | 2008 - 2011 | 8527 | 26.6% | 10498 | 25.7% | 10309 | 26.1% |
|  | 2012 - 2016 | 9325 | 29.0% | 14456 | 35.3% | 14989 | 38.0% |
| Practice Region | North East | 1118 | 3.5% | 1383 | 3.4% | 1421 | 3.6% |
|  | North West | 4086 | 12.7% | 4923 | 12.0% | 5131 | 13.0% |
|  | Yorkshire & The Humber | 1482 | 4.6% | 2089 | 5.1% | 1950 | 4.9% |
|  | East Midlands | 919 | 2.9% | 1451 | 3.5% | 1394 | 3.5% |
|  | West Midlands | 4649 | 14.5% | 5609 | 13.7% | 5392 | 13.7% |
|  | East of England | 3123 | 9.7% | 4321 | 10.6% | 4132 | 10.5% |
|  | South West | 5561 | 17.3% | 7531 | 18.4% | 7104 | 18.0% |
|  | South Central | 4142 | 12.9% | 5245 | 12.8% | 4649 | 11.8% |
|  | London | 2773 | 8.6% | 3577 | 8.7% | 3689 | 9.4% |
|  | South East Coast | 3904 | 12.2% | 4743 | 11.6% | 4541 | 11.5% |
|  | Missing | 359 | 1.1% | 43 | 0.1% | 40 | 0.1% |
| IMD | 1 - least deprived | 5864 | 18.3% | 7342 | 17.9% | 6913 | 17.5% |
|  | 2 | 5911 | 18.4% | 7482 | 18.3% | 7138 | 18.1% |
|  | 3 | 6591 | 20.5% | 8368 | 20.5% | 8041 | 20.4% |
|  | 4 | 6876 | 21.4% | 9104 | 22.3% | 8787 | 22.3% |
|  | 5 - most deprived | 6555 | 20.4% | 8619 | 21.1% | 8564 | 21.7% |
|  | Missing | 319 | 1.0% | 0 | 0.0% | 0 | 0.0% |
| Ethnicity | Asian | 307 | 1.0% | 396 | 1.0% | 389 | 1.0% |
|  | Black | 687 | 2.1% | 842 | 2.1% | 823 | 2.1% |
|  | Mixed | 93 | 0.3% | 108 | 0.3% | 94 | 0.2% |
|  | Other | 140 | 0.4% | 195 | 0.5% | 189 | 0.5% |
|  | Unknown | 1672 | 5.2% | 1690 | 4.1% | 1109 | 2.8% |
|  | White | 29217 | 91.0% | 37684 | 92.1% | 36839 | 93.4% |

### 3.2 Accuracy and completeness estimates

Among the 51,487 patients, 32,116 (62.4%) were identified through CPRD, 40,915 (79.5%) from NCRAS, and 39,443 (76.6%) from HES (Fig. 2). Of these patients 22,161 (43%) had their diagnosis commonly documented in all the data sources, while 38,826 (75.4%) had it documented in at least two. 12,661 (24.6%) of the cases were identified in only one source with the highest being in HES (12.4%) and the least in CPRD (4.4%).

**Fig. 2.**
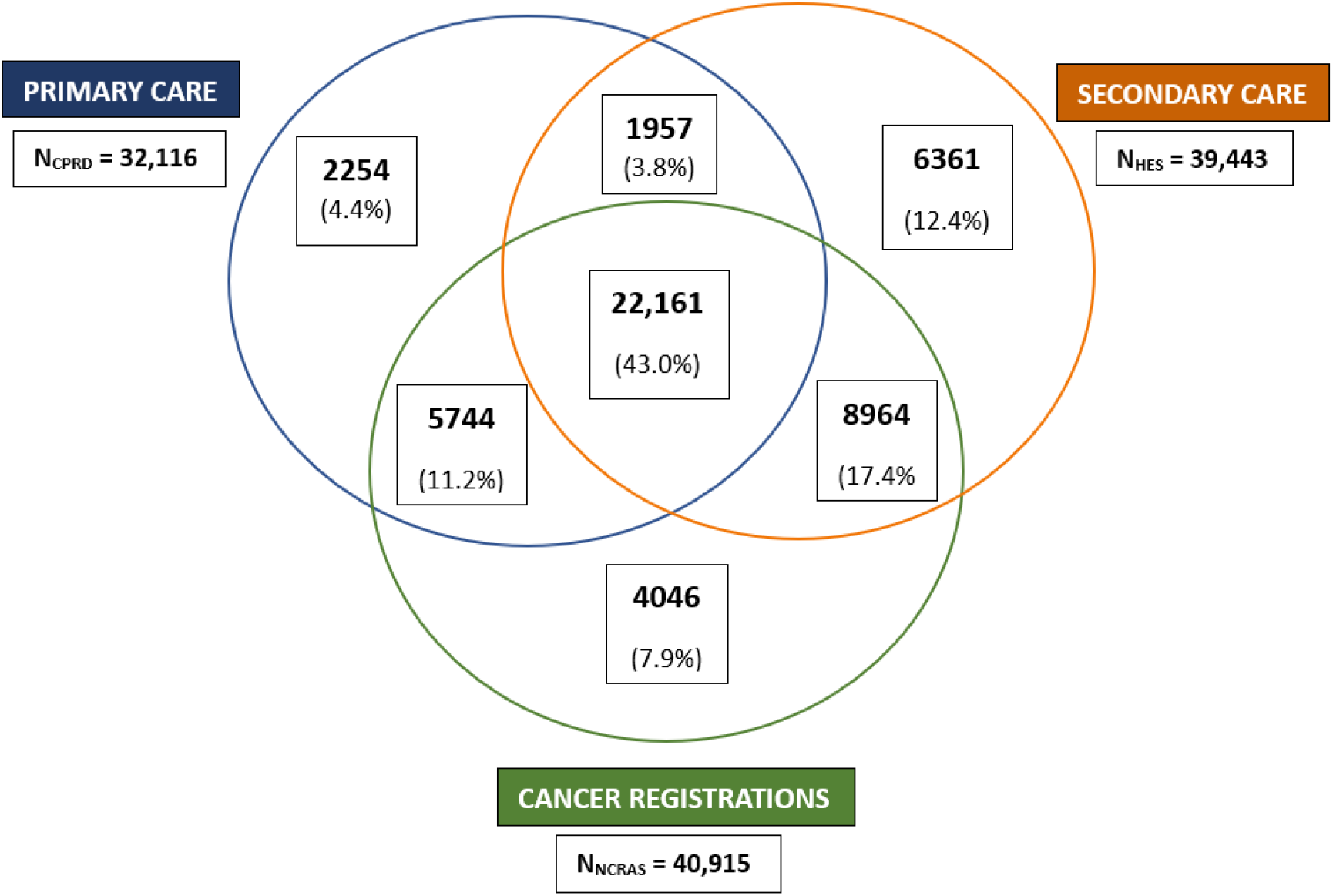
Comparison of cases identified from each data source

CPRD-NCRAS demonstrated higher accuracy (86.9%) than CPRD-HES (75.1%) (Table 2). Accuracy was lowest for patients aged ≥80 years, with 78.6% for CPRD-NCRAS and 73.2% for CPRD-HES. Over time, accuracy of CPRD-NCRAS improved to 91.4% during 2012-2016, while CPRD-HES declined to 69.0%. IMD quintile 5 exhibited high accuracy with both NCRAS (89.5%) and HES (79.6%). The ‘mixed’ ethnic group showed higher accuracy in CPRD-NCRAS (92.5%) but the lowest accuracy in CPRD-HES (68.8%).

**Table 2.** Accuracy estimates of prostate cancer diagnoses recorded in CPRD compared to NCRAS, HES and combined NCRAS-HES data.

| Accuracy of prostate cancer diagnoses in CPRD |  | Cases in CPRD (N) | Comparison with NCRAS |  | Comparison with HES |  | Comparison with NCRAS and/or HES |  |
| --- | --- | --- | --- | --- | --- | --- | --- | --- |
|  |  |  | Cases in CPRD and NCRAS (N) | Accuracy (%) | Cases in CPRD and HES (N) | Accuracy (%) | Cases in CPRD and NCRAS and/or HES (N) | Accuracy (%) |
| **Overall accuracy estimates** |  | 32,116 | 27,905 | 86.9% | 24,118 | 75.1% | 29,862 | 93.0% |
| **Age at diagnosis (years)** | < 60 | 1004 | 885 | 88.1% | 773 | 77.0% | 933 | 92.9% |
|  | 60 - 69 | 9812 | 8884 | 90.5% | 7555 | 77.0% | 9272 | 94.5% |
|  | 70 - 79 | 13439 | 11957 | 89.0% | 10038 | 74.7% | 12665 | 94.2% |
|  | ≥ 80 | 7861 | 6179 | 78.6% | 5752 | 73.2% | 6992 | 88.9% |
| **Year of diagnosis** | 2000 - 2003 | 6149 | 4795 | 78.0% | 4700 | 76.4% | 5462 | 88.8% |
|  | 2004 - 2007 | 8115 | 6901 | 85.0% | 6381 | 78.6% | 7527 | 92.8% |
|  | 2008 - 2011 | 8527 | 7690 | 90.2% | 6606 | 77.5% | 8093 | 94.9% |
|  | 2012 - 2016 | 9325 | 8519 | 91.4% | 6431 | 69.0% | 8780 | 94.2% |
| **IMD** | 1 - least deprived | 5864 | 5080 | 86.6% | 4267 | 72.8% | 5454 | 93.0% |
|  | 2 | 5911 | 5164 | 87.4% | 4380 | 74.1% | 5525 | 93.5% |
|  | 3 | 6591 | 5764 | 87.5% | 4973 | 75.5% | 6199 | 94.1% |
|  | 4 | 6876 | 6027 | 87.7% | 5279 | 76.8% | 6460 | 93.9% |
|  | 5 - most deprived | 6555 | 5870 | 89.5% | 5219 | 79.6% | 6224 | 95.0% |
| **Ethnicity** | Asian | 307 | 270 | 87.9% | 232 | 75.6% | 288 | 93.8% |
|  | Black | 687 | 600 | 87.3% | 528 | 76.9% | 648 | 94.3% |
|  | Mixed | 93 | 86 | 92.5% | 64 | 68.8% | 88 | 94.6% |
|  | Other | 140 | 122 | 87.1% | 104 | 74.3% | 135 | 96.4% |
|  | White | 29217 | 25797 | 88.3% | 22630 | 77.5% | 27602 | 94.5% |

Completeness estimates were 68.5% for CPRD-NCRAS and 61.1% for CPRD-HES (Table 3). They decreased with increasing age and was lowest during 2012-2016 (59.1%) for CPRD-NCRAS and during 2000-2003 (53.4%) for CPRD-HES. Completeness of IMD 1-3 was higher than of IMD 5 with both sources. The ‘mixed’ ethnic group showed the highest completeness in both CPRD-NCRAS (79.6%) and CPRD-HES (68.1%). Overall accuracy for CPRD, NCRAS, and HES were estimated as 93.0%, 90.1%, and 83.9% respectively, while completeness estimates were 60.7%, 77.7%, and 73.3% respectively.

**Table 3.**
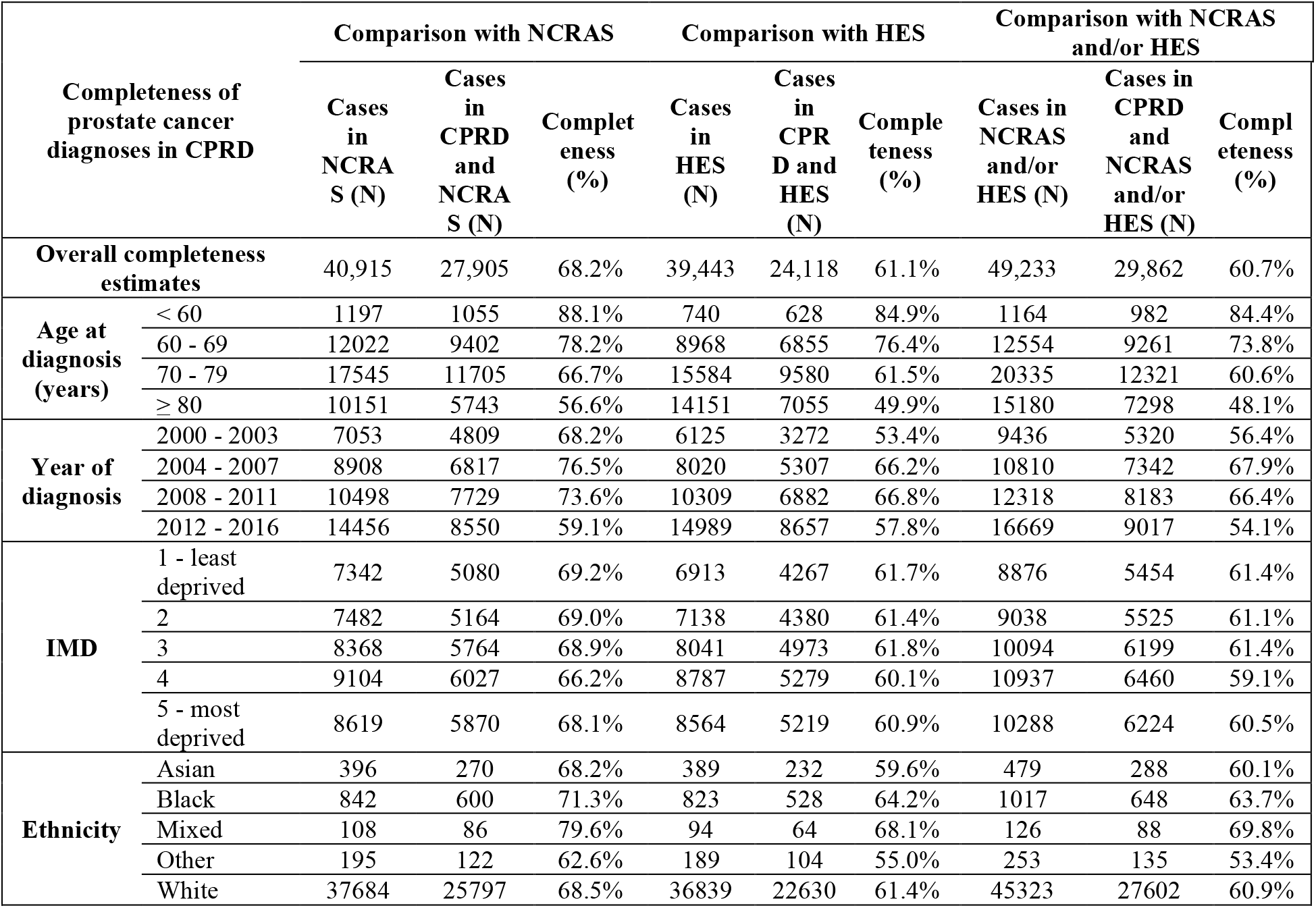
Completeness estimates of prostate cancer diagnoses recorded in CPRD compared to NCRAS, HES and combined NCRAS-HES data.

| Completeness of prostate cancer diagnoses in CPRD |  | Comparison with NCRAS |  |  | Comparison with HES |  |  | Comparison with NCRAS and/or HES |  |  |
| --- | --- | --- | --- | --- | --- | --- | --- | --- | --- | --- |
|  |  | Cases in NCRAS (N) | Cases in CPRD and NCRAS (N) | Completeness (%) | Cases in HES (N) | Cases in CPRD and HES (N) | Completeness (%) | Cases in NCRAS and/or HES (N) | Cases in CPRD and NCRAS and/or HES (N) | Completeness (%) |
| Overall completeness estimates |  | 40,915 | 27,905 | 68.2% | 39,443 | 24,118 | 61.1% | 49,233 | 29,862 | 60.7% |
| Age at diagnosis (years) | < 60 | 1197 | 1055 | 88.1% | 740 | 628 | 84.9% | 1164 | 982 | 84.4% |
|  | 60 - 69 | 12022 | 9402 | 78.2% | 8968 | 6855 | 76.4% | 12554 | 9261 | 73.8% |
|  | 70 - 79 | 17545 | 11705 | 66.7% | 15584 | 9580 | 61.5% | 20335 | 12321 | 60.6% |
|  | ≥ 80 | 10151 | 5743 | 56.6% | 14151 | 7055 | 49.9% | 15180 | 7298 | 48.1% |
| Year of diagnosis | 2000 - 2003 | 7053 | 4809 | 68.2% | 6125 | 3272 | 53.4% | 9436 | 5320 | 56.4% |
|  | 2004 - 2007 | 8908 | 6817 | 76.5% | 8020 | 5307 | 66.2% | 10810 | 7342 | 67.9% |
|  | 2008 - 2011 | 10498 | 7729 | 73.6% | 10309 | 6882 | 66.8% | 12318 | 8183 | 66.4% |
|  | 2012 - 2016 | 14456 | 8550 | 59.1% | 14989 | 8657 | 57.8% | 16669 | 9017 | 54.1% |
| IMD | 1 - least deprived | 7342 | 5080 | 69.2% | 6913 | 4267 | 61.7% | 8876 | 5454 | 61.4% |
|  | 2 | 7482 | 5164 | 69.0% | 7138 | 4380 | 61.4% | 9038 | 5525 | 61.1% |
|  | 3 | 8368 | 5764 | 68.9% | 8041 | 4973 | 61.8% | 10094 | 6199 | 61.4% |
|  | 4 | 9104 | 6027 | 66.2% | 8787 | 5279 | 60.1% | 10937 | 6460 | 59.1% |
|  | 5 - most deprived | 8619 | 5870 | 68.1% | 8564 | 5219 | 60.9% | 10288 | 6224 | 60.5% |
| Ethnicity | Asian | 396 | 270 | 68.2% | 389 | 232 | 59.6% | 479 | 288 | 60.1% |
|  | Black | 842 | 600 | 71.3% | 823 | 528 | 64.2% | 1017 | 648 | 63.7% |
|  | Mixed | 108 | 86 | 79.6% | 94 | 64 | 68.1% | 126 | 88 | 69.8% |
|  | Other | 195 | 122 | 62.6% | 189 | 104 | 55.0% | 253 | 135 | 53.4% |
|  | White | 37684 | 25797 | 68.5% | 36839 | 22630 | 61.4% | 45323 | 27602 | 60.9% |

### 3.3 Discrepancies in diagnosis dates

In CPRD-NCRAS 25,287 (90.6%) were recorded within 6 months whereas in CPRD-HES it was only 14,763 (61.2%), suggesting better timing concordance with NCRAS (Supplementary_information-S4). Agreement on diagnosis dates improved over time, particularly after 2004, with both NCRAS and HES. Discrepancies were quantified in weeks with positive x-axis values representing later dates in CPRD and negative values representing earlier dates (Fig. 3). In CPRD-NCRAS 2,809 (11.1%) (Fig. 3a) and in CPRD-HES, 1,241 (8.4%) (Fig. 3b) shared the same date. Concordance in diagnosis dates was slightly higher in CPRD-NCRAS than in CPRD-HES. Later diagnosis dates in CPRD were observed for 77.2% of the patients compared to NCRAS and 53.3% compared to HES. The median difference between the diagnosis dates was 2 weeks later in CPRD (IQR 1 to 4 weeks) compared to NCRAS and 1 week later in CPRD (IQR -7 to 3 weeks) compared to HES. There were peaks at 2 weeks, indicating 4,877 (19.3%) cases in NCRAS and 2,015 (13.6%) cases in HES were recorded 2 weeks later in CPRD. In HES, another distinct peak showed that 1,935 cases (13.1%) were recorded at least 3 months earlier in CPRD compared to HES.

**Fig. 3.**
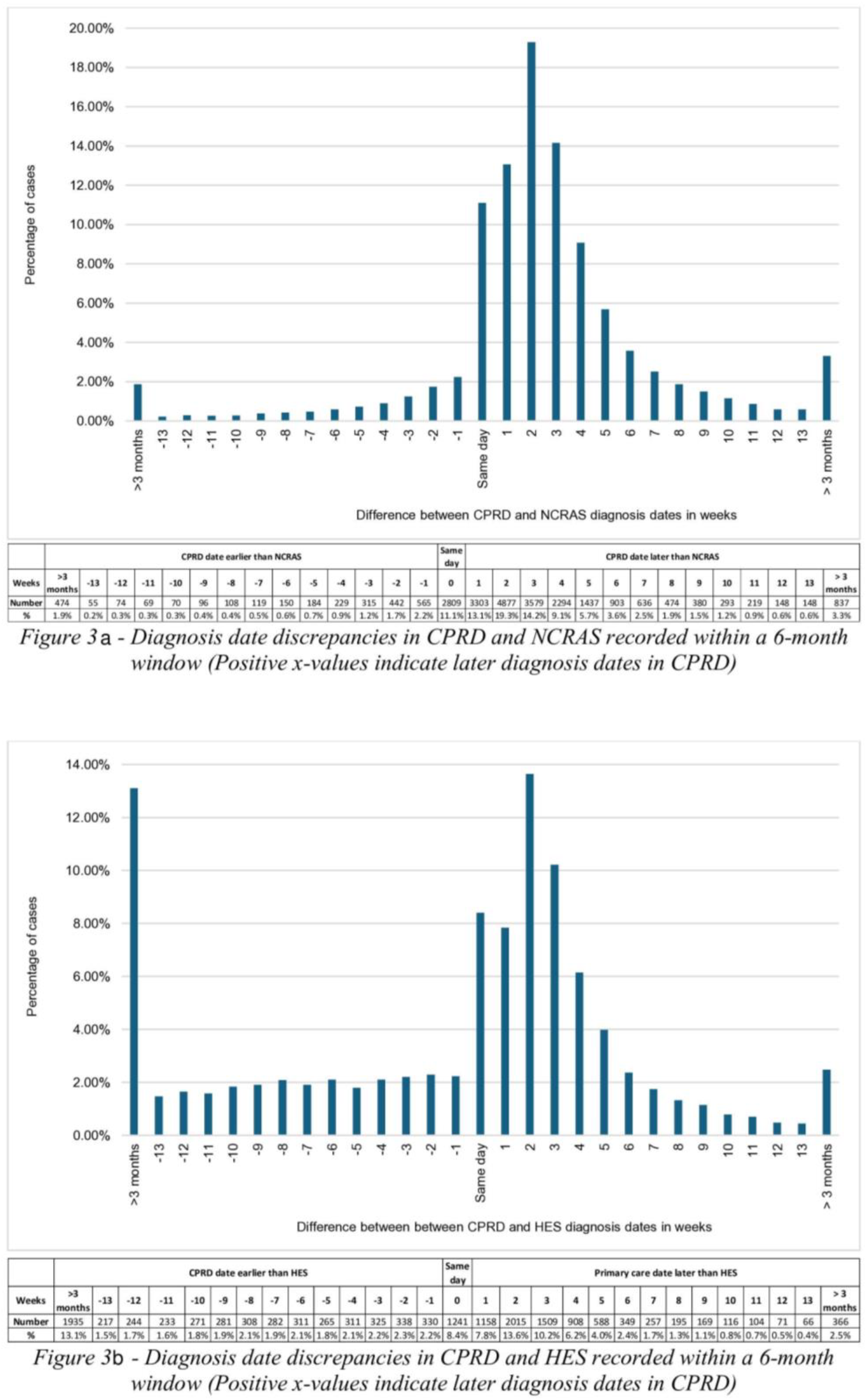
Diagnosis date discrepancies in CPRD with the dates in NCRAS and HES

### 3.4 Comparison of CPRD GOLD and Aurum

CPRD recorded PCa diagnoses for 57.4% in GOLD and 67.9% in Aurum (Fig. 4). Both CPRD-NCRAS had diagnoses recorded for 49.6% in GOLD and 59.4% in Aurum, while CPRD-HES had 42.5% in GOLD and 51.7% in Aurum. In GOLD 53.1% and in Aurum 64% had diagnoses recorded in CPRD and at least one other source while 39% in GOLD and 48% in Aurum had diagnoses present in all three sources. In GOLD 42.6 % of patients and in Aurum 32.1% had diagnoses in NCRAS or HES but were not recorded in CPRD.

**Fig. 4.**
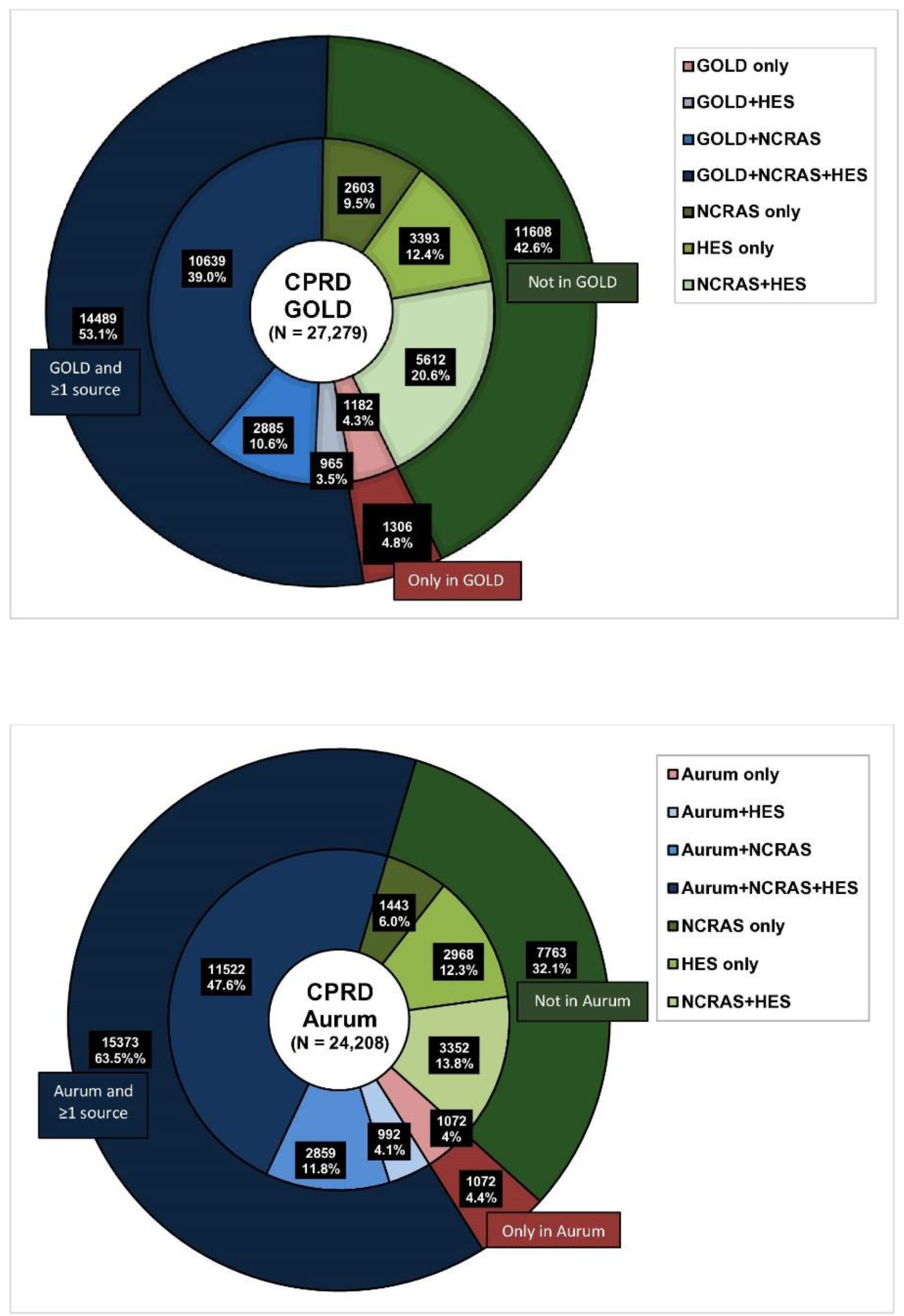
Prostate cancer diagnoses identified by source in CPRD GOLD vs CPRD Aurum

Of the 51,487 patients, 8964 (17.4%) had PCa diagnoses recorded in both NCRAS and HES but not in CPRD. We explored these patient records within 2 years of their NCRAS diagnosis date to identify potential reasons for the absence of PCa diagnosis codes in CPRD (Supplementary_information-S5 and S6). Aurum (37.4%) compared to GOLD (62.6%) had a lower proportion of missing CPRD diagnoses. Among the missing cases, 6.5% were identified as PCa diagnoses in CPRD but were not recorded during the study period. In Aurum 34.2% and in GOLD 19.2% of the patients had non-diagnostic codes recorded that were suggestive of PCa. In this study, we only considered malignant PCa diagnoses but 13.7% of the patients had non-malignant PCa codes in CPRD. Around 8% in both GOLD and Aurum had diagnosis codes for other prostate conditions such as prostatism often used interchangeably with benign prostatic hyperplasia. In both cohorts, 48% of patients had administrative codes (e.g., letter from specialist) that likely contained diagnostic data but were not documented in CPRD.

## 4. Discussion

### 4.1 Overview

This study evaluated the quality of primary care PCa diagnosis recording based on accuracy, completeness and timing of diagnosis using major English national datasets. We expected PCa diagnoses to be captured within linked data sources, considering the pathway involving hospital treatment, cancer registry reporting, and follow-up care by GPs and specialists.

NCRAS identified the majority of cases, while CPRD identified the least. HES had an older patient cohort than CPRD and NCRAS. CPRD had more patients from the least deprived areas but fewer from the most deprived areas compared to linked sources. CPRD showed higher accuracy but lower completeness than other sources. Notably, 8% of CPRD cases were absent in NCRAS, about 20% in HES, while 28.6% of NCRAS or HES cases were missing in CPRD.

Concordance of diagnosis dates was better in CPRD-NCRAS (90.6% within 6 months) than in CPRD-HES (61.2%), improving over time, especially after 2004, likely due to initiatives like the Quality and Outcomes Framework (QOF). CPRD recorded diagnoses later than NCRAS and HES, with median delays of 2 weeks and 1 week, respectively. Aurum showed better accuracy (64%) and completeness (66%) compared to GOLD (53% and 56%). Additionally, 8964 (17.4%) patients had diagnoses in NCRAS and HES without corresponding codes in CPRD. Exploring these CPRD records revealed non-diagnostic PCa codes, non-malignant PCa codes, conditions like prostatism, and vague administrative codes.

### 4.2 Comparison with previous studies

Previous studies on cancer recording quality in CPRD, particularly GOLD, have shown varied accuracy and completeness by cancer site and year (9,27–30). Margulis et al. (9) observed higher completeness for PCa in GOLD during 2004-2012, though their cohort only included patients treated for overactive bladder, limiting generalisability. Boggon et al. (30) and Williams et al. (31) reported higher accuracy for PCa in CPRD compared to NCRAS, consistent with our findings. Arhi et al. (29) reported 10% of the cases in CPRD or HES without corresponding diagnoses in NCRAS similar to our findings. Missing cases in NCRAS might be due to differences in diagnosis and coding, diagnoses from private healthcare or outside England, or data linkage errors. Strongman et al. (31) developed a gold standard algorithm using multiple data sources, identifying cases in GOLD, HES, or ONS that were missing in NCRAS, highlighting the importance of data linkage.

Discrepancies in diagnosis dates, with more cases having later dates in CPRD than NCRAS and HES, align with previous research (21, 25, 46). Peaks at two weeks suggest the standard time for secondary care reports/letters to be written, transmitted to primary care, and then coded in the patient record. Arhi et al. (29) noted two peaks in CPRD with HES suggesting the first peak, occurring over 3 months earlier in CPRD reflected the extended period between neo-adjuvant treatment and surgery while the second peak indicated earlier HES diagnoses, potentially following resection, or an inpatient procedure. Studies have reported that the use of HES alone over-represents older patients similar to our findings (29,31,32).

Differences in data sources, study periods, diagnostic codes, and population criteria may explain discrepancies between prior studies and our findings. Our results, along with previous research, affirm the quality of PCa diagnoses in CPRD for observational research compared to linked NCRAS and HES data, highlighting the potential for improved PCa diagnosis data quality through data linkage.

### 4.3 Strengths and limitations

This study, to our knowledge, is the first to assess the quality of PCa diagnosis recording in CPRD by combining both GOLD and Aurum datasets. Estimating accuracy and completeness by patient’s age, diagnosis year, IMD and ethnicity helped identify potential disparities in data quality across demographic groups. The comparison between GOLD and Aurum supports researchers in selecting data sources for PCa research. Investigating why patients had diagnostic codes in NCRAS and/or HES but not in CPRD highlighted challenges in case identification and the limitations of coded data. The study emphasised the importance of data linkage in CPRD due to the lack of a definitive gold standard for identifying all PCa cases.

Differences in databases, GP software, and coding systems may introduce biases impacting study outcomes (33). Discrepancies in coding dictionaries are a potential limitation, although the use of predefined codes and team reviews aimed to mitigate these issues. The exclusive use of PCa diagnostic codes without combining other domains (e.g., test/morphology/treatment) may have affected case identification (34). Reliance on structured data might miss PCa cases where diagnostic information exists in clinical notes. While the study compared the quality of GOLD to Aurum, it did not separately focus on each, as this would have required extensive additional analyses and hindered interpretation. The applicability of the results might be limited due to the study period and post-COVID-19 changes, and the age group may restrict generalisability albeit the prevalence of PCa among older patients over 50 in the UK.

### 4.4 Implications and future steps

Our study underscores the importance of integrating multiple data sources for PCa research. While CPRD data shows higher accuracy, it has lower completeness. Conversely, NCRAS captures more cases but may have lower accuracy. Thus, combining CPRD with linked sources can improve the quality of PCa diagnosis. However, no single data source is a definitive gold standard, and researchers must account for the limitations of each resource and the practical implications of linked data use. Future research should investigate reasons for unconfirmed or missing cases, establish a cohort based on these findings, compare patient characteristics between GOLD and Aurum, and examine the impact of global events like the pandemic on PCa recording.

## 5. Conclusions

In conclusion, the study demonstrates high accuracy for PCa recording in CPRD compared to NCRAS and HES, affirming its suitability for research purposes. However, for complete case capture, linking to HES or NCRAS is recommended. Researchers must address inherent limitations within each data source, tailor approaches to study requirements, and mitigate disparities in recording, timing, and patient characteristics to ensure research validity.

## Data Availability

The data utilised in this study were obtained from the CPRD, facilitated by the UK MHRA. However, the authors' license for using these data does not permit the sharing of raw data with third parties. For information regarding access to CPRD data, interested parties may refer to the following link: Research applications | CPRD.

## Statements and Declarations

### Authors’ contributions

GS, AL, and EF contributed to the conceptualisation of the study; GS carried out the main analysis and wrote the manuscript; AL, EF, JA and SM provided advice on the methodology and completed the subsequent revisions and editing of the manuscript; MC and PF provided advice on the data. All authors commented on successive drafts, and read and approved the final manuscript.

### Ethics statement

The study was approved by the Medicines and Healthcare Products Regulatory Agency (MHRA) Independent Scientific Advisory Committee (protocol number 19_050R).

### Footnotes

## Acknowledgements

JA & EF receive funding from the National Institute for Health and Care Research (NIHR) Applied Research Collaboration Kent, Surrey, Sussex. The views expressed are those of the author(s) and not necessarily those of the NHS, the NIHR or the Department of Health and Social Care.

## Provenance and peer review

Not commissioned, externally peer-reviewed.

## Additional Information

Supplementary material available in ***Supplementary Information (SI)*.*pdf***

Correspondence and requests for materials should be addressed to Gayasha Somathilake.

Software developed for data extraction and analysis with the code lists used to define variables in this study will be made openly accessible for review and reuse on GitHub (https://github.com/).

