## Supplementary Information (SI) for "Evaluating the quality of prostate cancer diagnosis recording in routinely collected primary care data for observational research: A study using multiple linked English electronic health records databases"

### 1. Code list for prostate cancer (S1)

The Read and ICD-10 codes used to define and extract prostate cancer diagnoses from the data sources are included below along with the corresponding med codes used for GOLD and Aurum primary care databases.

| Coding Terminology | Code | Term | GOLD medcode | Aurum medcode |
|---|---|---|---|---|
| Read codes | B46..00 | Malignant neoplasm of prostate | 780 | 722361000006115 |
| | 4M01.00 | Gleason prostate grade 5-7 (medium) | 18612 | 1488634012 |
| | 4M02.00 | Gleason prostate grade 8-10 (high) | 26081 | 1490807013 |
| | HNG0200 | [RFC] Cancer of the prostate | - | 906971000006110 |
| ICD-10 codes | C61 | Malignant neoplasm of prostate | | |

### 2. Timelines of each data source (S2)

The time periods for data availability in each of the data source are depicted below and the study period was chosen when the data was concurrently available.

***CPRD*** *– Clinical Practice Research Datalink,* ***NCRAS*** *- National Cancer Registration and Analysis Service,* ***CR****- Cancer Registrations,* ***RTDS*** *– Radiotherapy Data Set,* ***SACT*** *- Systemic Anti-Cancer Therapy,* ***HES*** *- Hospital Episode Statistics,* ***APC*** *– Admitted Patient Care,* ***OP*** *– Outpatient*

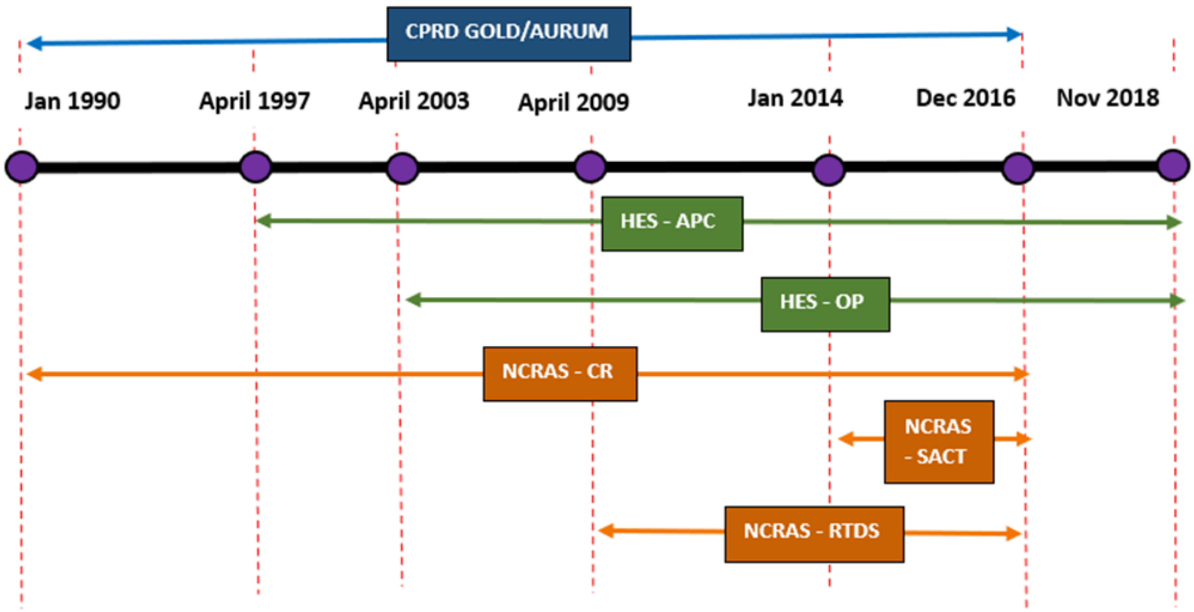

3. **Reporting guidelines (S3)**

The reporting guidelines based on the STROBE RECORD (The REporting of studies Conducted using Observational Routinely-collected health Data) statement was followed in this study and the checklist used is provided.

**The RECORD statement – checklist of items, extended from the STROBE statement, that should be reported in observational studies using routinely collected health data.**

| | Item No. | RECORD items | Location in manuscript where items are reported |
|---|---|---|---|
| **Title and abstract** | | | |
| Title and abstract | 1 | RECORD 1.1: The type of data used should be specified in the title or abstract. When possible, the names of the databases used should be included.<br><br>RECORD 1.2: If applicable, the geographic region and timeframe within which the study took place should be reported in the title or abstract.<br><br>RECORD 1.3: If linkage between databases was conducted for the study, this should be clearly stated in the title or abstract. | Abstract – Background and Methods (Page 1) |
| **Introduction** | | | |
| Background rationale | 2 | Explain the scientific background and rationale for the investigation being reported | Background - Paragraphs 1, 2 and 3 (Pages 2-3) |
| Objectives | 3 | State the specific objectives, including any prespecified hypotheses | |
| **Methods** | | | |
| Study Design | 4 | Present key elements of study design early in the paper | Methods – Data sources, Patient selection (Pages 4-6) |
| Setting | 5 | Describe the setting, locations, and relevant dates, including periods of recruitment, exposure, follow-up, and data collection | |
| Participants | 6 | RECORD 6.1: The methods of study population selection (such as codes or algorithms used to identify subjects) should be listed in detail. If this is not possible, an explanation should be provided.<br><br>RECORD 6.2: Any validation studies of the codes or algorithms used to select the population should be referenced. If validation was conducted for this study and not published | Methods – Patient selection (Page 5) and Supplementary_ information - S1<br><br>Results – Study cohort – Figure 1 (Page 8) |

| | | | |
|---|---|---|---|
| | | elsewhere, detailed methods and results should be provided.<br><br>RECORD 6.3: If the study involved linkage of databases, consider use of a flow diagram or other graphical display to demonstrate the data linkage process, including the number of individuals with linked data at each stage. | |
| Variables | 7 | RECORD 7.1: A complete list of codes and algorithms used to classify exposures, outcomes, confounders, and effect modifiers should be provided. If these cannot be reported, an explanation should be provided. | Methods – Patient characteristics (Page 6) |
| Data sources/ measurement | 8 | For each variable of interest, give sources of data and details of methods of assessment (measurement).<br>Describe comparability of assessment methods if there is more than one group | |
| Bias | 9 | Describe any efforts to address potential sources of bias | Methods – Patient selection (Pages 5-6) |
| Study size | 10 | Explain how the study size was arrived at | Results – Study cohort – Figure 1 (Page 8) |
| Quantitative variables | 11 | Explain how quantitative variables were handled in the analyses. If applicable, describe which groupings were chosen, and why | Methods – Patient characteristics (Page 6) |
| Statistical methods | 12 | (a) Describe all statistical methods, including those used to control for confounding<br>(b) Describe any methods used to examine subgroups and interactions<br>(c) Explain how missing data were addressed<br>(d) *Cohort study* - If applicable, explain how loss to follow-up was addressed<br>*Case-control study* - If applicable, explain how matching of cases and controls was addressed<br>*Cross-sectional study* - If applicable, describe analytical methods taking account of sampling strategy<br>(e) Describe any sensitivity analyses | Methods – Data analysis (Pages 6-7) |
| Data access and cleaning methods | | RECORD 12.1: Authors should describe the extent to which the investigators had access to the database population used to create the study population.<br><br>RECORD 12.2: Authors should provide information on the data cleaning methods used in the study. | Methods – Data sources, Patient selection<br>(Pages 4-6) |

| | | | |
|---|---|---|---|
| Linkage | | RECORD 12.3: State whether the study included person-level, institutional-level, or other data linkage across two or more databases. The methods of linkage and methods of linkage quality evaluation should be provided. | |
| **Results** | | | |
| Participants | 13 | RECORD 13.1: Describe in detail the selection of the persons included in the study (*i.e.*, study population selection) including filtering based on data quality, data availability and linkage. The selection of included persons can be described in the text and/or by means of the study flow diagram. | Results – Study cohort – Figure 1 (Pages 7-8) |
| Descriptive data | 14 | (a) Give characteristics of study participants (*e.g.*, demographic, clinical, social) and information on exposures and potential confounders<br>(b) Indicate the number of participants with missing data for each variable of interest<br>(c) *Cohort study* - summarise follow-up time (*e.g.*, average and total amount) | Results – Study cohort – Table 1 (Page 9) |
| Outcome data | 15 | *Cohort study* - Report numbers of outcome events or summary measures over time<br>*Case-control study* - Report numbers in each exposure category, or summary measures of exposure<br>*Cross-sectional study* - Report numbers of outcome events or summary measures | Results (Pages 10-16) |
| Main results | 16 | (a) Give unadjusted estimates and, if applicable, confounder-adjusted estimates and their precision (e.g., 95% confidence interval). Make clear which confounders were adjusted for and why they were included<br>(b) Report category boundaries when continuous variables were categorized<br>(c) If relevant, consider translating estimates of relative risk into absolute risk for a meaningful time period | |
| Other analyses | 17 | Report other analyses done—e.g., analyses of subgroups and interactions, and sensitivity analyses | |
| **Discussion** | | | |
| Key results | 18 | Summarise key results with reference to study objectives | Discussion – Overview (Page 17) |
| Limitations | 19 | RECORD 19.1: Discuss the implications of using data that were not created or collected to answer the specific research question(s). Include discussion of misclassification bias, unmeasured confounding, missing data, and changing | Discussion – Strengths and limitations (Pages 20-21) |

| | | | |
|---|---|---|---|
| | | eligibility over time, as they pertain to the study being reported. | |
| Interpretation | 20 | Give a cautious overall interpretation of results considering objectives, limitations, multiplicity of analyses, results from similar studies, and other relevant evidence | Discussion – Comparison with previous studies, Implications and future steps (Pages 18,19,21) |
| Generalisability | 21 | Discuss the generalisability (external validity) of the study results | Conclusion (Page 21) |
| **Other information** | | | |
| Funding | 22 | Give the source of funding and the role of the funders for the present study and, if applicable, for the original study on which the present article is based | Footnotes (Page 29) |
| Accessibility of protocol, raw data, and programming code | | RECORD 22.1: Authors should provide information on how to access any supplemental information such as the study protocol, raw data, or programming code. | |

*Reference: Benchimol EI, Smeeth L, Guttmann A, Harron K, Moher D, Petersen I, Sørensen HT, von Elm E, Langan SM, the RECORD Working Committee. The REporting of studies Conducted using Observational Routinely-collected health Data (RECORD) Statement. *PLoS Medicine* 2015; in press.

*Checklist is protected under Creative Commons Attribution (CC BY) license.

### 4. Timing variations in prostate cancer diagnosis recording (S4)

Variations in the prostate cancer diagnosis recording dates in CPRD (primary care) compared to NCRAS and HES are assessed based on the calendar year.

| Primary care | | NCRAS | | | HES | | |
|---|---|---|---|---|---|---|---|
| Index year | Total number of cases in primary care (N , col %) | Within 6 months (N , row %) | Over 6 months (N , row %) | Total number of cases (N , col %) | Within 6 months (N , row %) | Over 6 months (N , row %) | Total number of cases (N , col %) |
| **2000** | 1257 (3.9%) | 724 (82.1%) | 158 (17.9%) | 882 (3.2%) | 491 (53.8%) | 421 (46.2%) | 912 (3.8%) |
| **2001** | 1476 (4.6%) | 927 (81.8%) | 206 (18.2%) | 1133 (4.1%) | 579 (51.8%) | 539 (48.2%) | 1118 (4.6%) |
| **2002** | 1627 (5.1%) | 1033 (80.1%) | 257 (19.9%) | 1290 (4.6%) | 647 (50.6%) | 631 (49.4%) | 1278 (5.3%) |
| **2003** | 1789 (5.6%) | 1220 (81.9%) | 270 (18.1%) | 1490 (5.3%) | 686 (49.3%) | 706 (50.7%) | 1392 (5.8%) |
| **2004** | 2002 (6.2%) | 1436 (86.1%) | 232 (13.9%) | 1668 (6.0%) | 845 (53.2%) | 742 (46.8%) | 1587 (6.6%) |
| **2005** | 1960 (6.1%) | 1484 (89.1%) | 181 (10.9%) | 1665 (6.0%) | 832 (54.2%) | 704 (45.8%) | 1536 (6.4%) |
| **2006** | 2111 (6.6%) | 1608 (89.0%) | 198 (11.0%) | 1806 (6.5%) | 918 (55.2%) | 745 (44.8%) | 1663 (6.9%) |
| **2007** | 2042 (6.4%) | 1620 (91.9%) | 142 (8.1%) | 1762 (6.3%) | 878 (55.0%) | 717 (45.0%) | 1595 (6.6%) |
| **2008** | 2081 (6.5%) | 1707 (91.7%) | 155 (8.3%) | 1862 (6.7%) | 944 (58.1%) | 680 (41.9%) | 1624 (6.7%) |
| **2009** | 2198 (6.8%) | 1812 (91.7%) | 165 (8.3%) | 1977 (7.1%) | 1037 (60.5%) | 678 (39.5%) | 1715 (7.1%) |
| **2010** | 2204 (6.9%) | 1865 (93.6%) | 128 (6.4%) | 1993 (7.1%) | 1071 (61.8%) | 661 (38.2%) | 1732 (7.2%) |
| **2011** | 2044 (6.4%) | 1731 (93.2%) | 127 (6.8%) | 1858 (6.7%) | 981 (63.9%) | 554 (36.1%) | 1535 (6.4%) |
| **2012** | 2113 (6.6%) | 1807 (94.7%) | 102 (5.3%) | 1909 (6.8%) | 1067 (67.2%) | 521 (32.8%) | 1588 (6.6%) |
| **2013** | 1966 (6.1%) | 1734 (94.7%) | 97 (5.3%) | 1831 (6.6%) | 998 (68.9%) | 450 (31.1%) | 1448 (6.0%) |
| **2014** | 1907 (5.9%) | 1665 (95.4%) | 80 (4.6%) | 1745 (6.3%) | 1020 (74.2%) | 355 (25.8%) | 1375 (5.7%) |
| **2015** | 1777 (5.5%) | 1553 (96.3%) | 59 (3.7%) | 1612 (5.8%) | 967 (82.7%) | 202 (17.3%) | 1169 (4.8%) |
| **2016** | 1562 (4.9%) | 1361 (95.7%) | 61 (4.3%) | 1422 (5.1%) | 802 (94.2%) | 49 (5.8%) | 851 (3.5%) |
| **Total** | 32116 | 25287 (90.6%) | 2618 (9.4%) | 27905 | 14763 (61.2%) | 9355 (38.8%) | 24118 |

### 5. List of the most frequent terms found for patients without prostate cancer diagnosis codes in CPRD (S5)

The patient records that were identified with prostate cancer diagnoses in NCRAS and/or HES but with no corresponding diagnoses in CPRD were further investigated. The below table displays the terms that were recorded in primary care instead of diagnosis codes.

| CPRD GOLD (N = 3526) | | CPRD Aurum (N = 2045) | |
|---|---|---|---|
| **Term** | **Frequency** | **Term** | **Frequency** |
| Telephone encounter | 1936 | Prostate specific antigen | 1015 |
| Patient reviewed | 1532 | Medication review | 674 |
| Seen in urology clinic | 1236 | Seen in urology clinic | 617 |
| Had a chat to patient | 1026 | Telephone encounter | 582 |
| Medication review | 976 | Referral letter | 552 |
| Letter from specialist | 808 | Administration | 545 |
| Administration | 761 | Medication review done | 515 |
| Administration NOS | 749 | Referred to urologist | 427 |
| Discharge summary | 724 | Carcinoma in situ of prostate | 369 |
| Medication requested | 596 | Discharge summary | 353 |
| Carcinoma in situ of prostate | 572 | Patient reviewed | 333 |
| Seen in hospital casualty | 554 | Referral for further care | 308 |
| Letter encounter | 508 | Notes summary on computer | 292 |
| Result | 498 | Letter encounter | 290 |
| Notes summary on computer | 485 | Scanned document | 282 |
| Seen in oncology clinic | 438 | Letter from specialist | 268 |
| Advice to patient - subject | 435 | Result | 265 |
| Cancer care review | 404 | Seen in hospital casualty | 261 |
| Letter from consultant | 347 | Cancer care review | 257 |
| Prostatism | 313 | Had a chat to patient | 251 |
| Discussion | 312 | Discharged from hospital | 246 |
| Discharged from hospital | 309 | Seen in oncology clinic | 230 |
| Seen in general surgery clinic | 305 | Letter from consultant | 224 |
| Letter received | 289 | Transurethral prostatectomy | 186 |
| Transurethral prostatectomy | 277 | Medication requested | 183 |
| Medication review done | 265 | Discharge summary report | 181 |
| Letter encounter from patient | 263 | [D]Raised PSA | 176 |
| Discharge summary report | 240 | Suspected UTI | 168 |
| Referral for further care | 215 | Administration NOS | 156 |
| Prostate specific antigen | 200 | Emergency hospital admission | 156 |
| Patient given advice | 199 | Prostatism | 154 |
| [D]Raised PSA | 191 | Letter encounter from patient | 102 |
| Diagnostic cystoscopy | 179 | Letter received | 99 |
| Emergency hospital admission | 172 | Seen in general surgery clinic | 89 |
| Incoming mail NOS | 171 | Diagnostic cystoscopy | 83 |
| Referred to urologist | 152 | Fax received | 75 |
| Suspected UTI | 150 | Suspected prostate cancer | 72 |
| Fax received | 137 | Incoming mail NOS | 69 |
| Suspected prostate cancer | 131 | Advice to patient - subject | 63 |
| Insertion of hormone implant | 114 | Patient given advice | 59 |
| Seen in radiology department | 114 | Seen in radiology department | 55 |
| Prostatectomy NEC | 113 | Gleason grading of prostate cancer | 48 |
| Radiotherapy NEC | 89 | Prostatectomy NEC | 47 |
| H/O: prostate cancer | 40 | Radical prostatectomy - unspecified excision of pelvic nodes | 31 |
| Gleason grading of prostate cancer | 29 | Discussion | 29 |

| | | | |
|---|---|---|---|
| Radical prostatectomy without pelvic node excision | 29 | Insertion of hormone implant | 23 |
| Brachytherapy | 25 | Gleason prostate grade 2-4 (low) | 20 |
| Open prostatectomy | 22 | Radiotherapy NEC | 20 |
| Chemotherapy | 16 | Retropubic prostatectomy | 20 |
| Retropubic prostatectomy | 14 | H/O: prostate cancer | 18 |
| Seen in radiotherapy clinic | 14 | Radiotherapy completed | 15 |
| Radical prostatectomy - unspecified excision of pelvic nodes | 11 | [V]Personal history of malignant neoplasm of prostate | 9 |
| Chemotherapy follow-up | 7 | Secondary malignant neoplasm of prostate | 7 |
| Radical prostatectomy with pelvic lymphadenectomy | 7 | Radical prostatectomy with pelvic lymphadenectomy | 8 |
| Radical prostatectomy with pelvic node sampling | 7 | Radical prostatectomy with pelvic node sampling | 5 |
| Secondary malignant neoplasm of prostate | 7 | Seen in radiotherapy clinic | 5 |
| Gleason prostate grade 2-4 (low) | 6 | Neoplasm of uncertain behaviour of prostate | 4 |
| Prostatectomy planned | 6 | Date chemotherapy completed | 3 |
| Adenoma of prostate | 6 | Chemotherapy started | 2 |
| Radiotherapy | 4 | Open prostatectomy | 2 |
| Iodine seed radiotherapy | 3 | Chemotherapy started | 2 |
| Active surveillance | 2 | Active surveillance | 1 |
| Date chemotherapy completed | 2 | Laparoscopic approach | 1 |
| Discharge from radiotherapy service | 2 | Oral chemotherapy | 1 |
| Intravesical install chemotherapeutic agent for malignancy | 2 | Prostatectomy planned | 1 |
| Neoplasm of uncertain behaviour of prostate | 2 | | |
| Pre-operative chemotherapy | 2 | | |
| Laparoscopic approach | 2 | | |
| Seen by radiotherapist | 2 | | |
| [V]Personal history of malignant neoplasm of prostate | 1 | | |
| Brachytherapy monitoring | 1 | | |
| Chemotherapy started | 1 | | |
| Discharge by radiotherapist | 1 | | |
| External radiotherapy NOS | 1 | | |
| H/O: chemotherapy | 1 | | |
| Intravenous chemotherapy | 1 | | |
| Oral chemotherapy | 1 | | |
| Progress of radiotherapy | 1 | | |
| Radiotherapy completed | 1 | | |
| Radiotherapy procedures | 1 | | |
| Radiotherapy progress NOS | 1 | | |

### 6. Summary table of the potential reasons for missing prostate cancer diagnosis codes in CPRD (S6)

The table summarises the potential reasons for lacking prostate cancer diagnosis codes in the CPRD records separately for GOLD and Aurum.

| **Reasons**<br>***(A patient may have more than one reason)*** | **CPRD GOLD (N=5612)** | **CPRD Aurum (N=3352)** | **CPRD (N=8964)** |
|---|---|---|---|
| Identified in CPRD as prostate cancer diagnosis but excluded (E.g. not during active follow-up) | 249 (4.4%) | 337 (10.1%) | 586 (6.5%) |
| Non-malignant prostate cancer codes recorded (E.g. in-situ, benign, history, Gleason 2-4) | 731 (13.0%) | 495 (14.8%) | 1226 (13.7%) |
| Codes suggestive of prostate cancer rather than a diagnosis are present (E.g. Treatments/ tests) | 1080 (19.2%) | 1147 (34.2%) | 2227 (24.8%) |
| Administrative codes present (E.g. “telephone encounter”, “letter from specialist”) | 2828 (50.4%) | 1484 (44.3%) | 4312 (48.1%) |
| Other prostate conditions (E.g. prostatitis) | 449 (8.0%) | 297 (8.9%) | 746 (8.3%) |
| Patients with no CPRD records during the period considered | 1837 (32.7%) | 970 (28.9%) | 2807 (31.3%) |
